# Analysis of SteraMist ionized hydrogen peroxide technology in the sterilization of N95 respirators and other PPE: a quality improvement study

**DOI:** 10.1101/2020.04.19.20069997

**Authors:** Avilash K. Cramer, Deborah Plana, Helen Yang, Mary M. Carmack, Enze Tian, Michael S. Sinha, David Krikorian, David Turner, Jinhan Mo, Ju Li, Rajiv Gupta, Heather Manning, Florence T. Bourgeois, Sherry H. Yu, Peter K. Sorger, Nicole R. LeBoeuf

**Author notes:** These authors contributed equally to this work (Avilash K. Cramer, Deborah Plana, Helen Yang). Corresponding author: Peter Sorger PhD, Otto Krayer Professor of Systems Biology, Warren Alpert Building 432, Program in Therapeutic Science, Harvard Medical School, 200 Longwood Avenue Boston, MA, 02115, (cc); (*not closely monitored during COVID pandemic*).

## Abstract

**Objective:** The COVID-19 pandemic has led to widespread shortages of personal protective equipment (PPE) for healthcare workers, including filtering facepiece respirators (FFRs) such as N95 masks. These masks are normally intended for single use, but their sterilization and subsequent reuse could substantially mitigate a world-wide shortage.

**Design:** Quality assurance.

**Setting:** A sealed environment chamber installed in the animal facility of an academic medical center.

**Interventions:** One to five sterilization cycles using ionized hydrogen peroxide (iHP), generated by SteraMist® equipment (TOMI; Frederick, MD).

**Main outcome measures:** Personal protective equipment, including five N95 mask models from three manufacturers, were evaluated for efficacy of sterilization following iHP treatment (measured with bacterial spores in standard biological indicator assemblies). Additionally, N95 masks were assessed for their ability to efficiently filter particles down to 0.3µm and for their ability to form an airtight seal using a quantitative fit test. Filtration efficiency was measured using ambient particulate matter at a university lab and an aerosolized NaCl challenge at a National Institute for Occupational Safety and Health (NIOSH) pre-certification laboratory.

**Results:** The data demonstrate that N95 masks sterilized using SteraMist iHP technology retain function up to five cycles, the maximum number tested to date. Some but not all PPE could also be sterilized using an iHP environmental chamber, but pre-treatment with a handheld iHP generator was required for semi-enclosed surfaces such as respirator hoses.

**Conclusions:** A typical iHP environment chamber with a volume of ~80 m^3^ can treat ~7000 masks per day, as well as other items of PPE, making this an effective approach for a busy medical center.

## Introduction

The COVID-19 pandemic has led to widespread shortages in personal protective equipment (PPE) for clinicians and first responders. Shortages in filtering facepiece respirators (FFRs) such as N95 “masks”, which are certified to filter 95% of airborne particles down to 0.3 µm, are particularly problematic because these normally single-use items are a mainstay of infection control. It has been widely reported that the US Department of Health and Human Services (HHS) anticipates a need for as many as 3.5 × 10^9^ N95 masks^1^ in 2020 for US use alone, but estimates of total available supply are far short of that number^2^. The consequent need for N95 mask sterilization and subsequent reuse is therefore likely to continue for the foreseeable future. The possibility that disposable N95 masks could be sterilized and reused was raised 15 years ago as a strategy to address shortages arising from medical emergencies,^3–5^ but with the exception of a single FDA-funded study by the Battelle Memorial Institute,^6^ little subsequent research was performed on the topic. Recently, in response to acute N95 mask shortages, multiple strategies for mask sterilization have been proposed and studied, including exposure to ultraviolet (UV) germicidal irradiation, vaporized hydrogen peroxide, moist heat, ethylene oxide, and gamma irradiation^7–14^. In this study, we evaluate a recently developed technology, ionized hydrogen peroxide (iHP), as a method for sterilizing N95 masks and other PPE.

Hydrogen peroxide (H_2_O_2_) is a powerful sterilizing agent that can be used on porous and other surfaces following vaporization or ionization to create a mist containing hydroxyl radicals. Such vaporized or ionized hydrogen peroxide methods (VHP/iHP) are widely used for environmental sterilization across multiple industries including food preparation, healthcare, and life sciences^14–19^. VHP/iHP methods can be used on a wider range of sensitive materials than high temperature methods (e.g. autoclaving) and are safer than ethylene oxide methods. Four distinct VHP/iHP-based H_2_O_2_ sterilization technologies that have been commercialized to date are shown in **Table 1**, each of which involves a different approach to generating and delivering the sterilant. In all cases microbial killing is achieved through the reaction of hydroxyl radicals with proteins, nucleic acids and other biomolecules in pathogens. Three VHP-based systems have received emergency use authorization (EUA) from the Food and Drug Administration (FDA) for N95 mask decontamination^20^, even though relatively limited peer-reviewed data is available^8^, particularly from non-commercial third parties. As a consequence, it is difficult for infection control teams in hospitals and other healthcare providers to evaluate these systems. The absence of data on the post-sterilization performance of different makes and models of N95 masks is also limiting. The Brigham and Women’s Hospital (BWH; Boston MA) Incident Command, which is involved in this study, currently has on hand over 30 models of N95 masks from three manufacturers.

**Table 1:** Commercial vaporization and ionization-based hydrogen peroxide sterilization technologies.

| Company | Products | Technology | Technology of delivery | Existing Use for N95 Sterilization |
| --- | --- | --- | --- | --- |
| Bioquell | Bioquell Clarus C;<br>Bioquell Z-2;<br>Bioquell ProteQ | **HPV**<br>*Hydrogen Peroxide Vapor* | 30-35% liquid H<sub>2</sub>O<sub>2</sub> vaporized and delivered into a chamber; saturated H<sub>2</sub>O<sub>2</sub> condenses on surfaces<sup>6,14,15,18</sup> | EUA granted (3/28/2020) to Batelle to use Bioquell as part of its Critical Care Decontamination System for up to 20 cycles of N95 mask reuse<sup>30</sup> |
| Steris Corporation | STERIS V-PRO 1 Plus, maX and maX2 Low Temperature Sterilization Systems | **VHP™**<br>*Vaporized Hydrogen Peroxide* | 30-35% liquid H<sub>2</sub>O<sub>2</sub> is vaporized and delivered into a dehumidified chamber; concentration is held below condensation point<sup>16</sup> | EUA granted (4/10/2020) to Steris Corporation for STERIS V-PRO 1 Plus, maX and maX2 Low Temperature Sterilization Systems for up to 10 cycles of single-user reuse<sup>31</sup> |
| Advanced Sterilization Products (ASP) | STERRAD 100S H<sub>2</sub>O<sub>2</sub> Gas Plasma Sterilizer; STERRAD NX H<sub>2</sub>O<sub>2</sub> Gas Plasma Sterilizer; STERRAD 100NX H<sub>2</sub>O<sub>2</sub> Gas Plasma Sterilizer | **HPGP**<br>*Hydrogen Peroxide Gas Plasma* | 58-60% liquid H<sub>2</sub>O<sub>2</sub> is vaporized into a chamber; radiation frequency energy is targeted into the chamber, exciting the H<sub>2</sub>O<sub>2</sub> to a low-temperature plasma state<sup>12,20</sup> | EUA granted (4/12/2020) to ASP for STERRAD 100S Cycle, STERRAD NX Standard Cycle, or STERRAD 100NX Express Cycle for up to two cycles in Tyvek pouching for single-user reuse<sup>29</sup> |
| TOMI | SteraMist Binary Ionization Technology (BIT) | **iHP™**<br>*ionized Hydrogen Peroxide* | 7.8% aqueous H<sub>2</sub>O<sub>2</sub> aerosolized. 0.05-3 µm droplets are pushed past a cold plasma field generated by two electrodes and ionized into hydroxyl radicals<sup>32</sup> | The topic of this study. Currently being investigated for sterilization of PPE for re-use in academic medical centers |

This study focuses on the use of iHP as an N95 mask sterilization method, specifically the SteraMist Binary Ionization Technology^®^ (BIT) from TOMI (Beverly Hills, CA). iHP was registered with the Environmental Protection Agency (EPA) in 2015 for use in health care, life sciences, food safety, and other settings (appearing on EPA lists G, H, K, L, and M). Most recently it was added to EPA List N: Disinfectants for Use Against SARS-CoV-2, for use on hard, nonporous surfaces^21^. The active ingredient in iHP is 7.8% aqueous H_2_O_2_ (an H_2_O_2_ concentration 5 times lower than used in commercial VHP systems), which is flowed past a plasma arc and dispersed into a treatment chamber as a mist of micron-sized liquid droplets. As it passes through the plasma, the hydrogen peroxide is ionized into reactive hydroxyl radicals, the active sterilant. iHP is commercially available in two implementations: a handheld sprayer device (“Surface Unit”) and an environmental unit (“Environment System”).

The environment system used in this study was installed at the Dana-Farber Cancer Institute (DFCI; Boston, MA) animal research facility for use in sterilizing incoming equipment and materials. Following cycles of sterilization, masks were tested for three critical features: (1) sterility, as measured by the inactivation of bacterial spores contained in biological indicators; (2) filtration efficiency, measured both by aerosolized 75 nm NaCl particles and by 0.3–1 µm ambient particulate matter; and (3) fit, using a PortaCount quantitative fit test apparatus. Multiple sterilization cycles were completed to assess mask durability. Sterilization of other PPE items such as face shields and hoods and hoses for Powered-Air Purifying Respirators (PAPRs) was also explored. Testing was performed at the DFCI, MIT and ICS Laboratories (a commercial laboratory accredited to perform testing to NIOSH/ISO/IEC standards).

## Methods

### Selection of N95 respirators and other PPE samples

A total of 73 N95 masks representing five models from three manufacturers (3M 1860, Kimberly-Clark [KC]/Halyard 46767 “duckbill,” Gerson 2130, 3M 8210, and 3M 9210/37021) were selected for testing as a representative sample of the N95 masks used in three local hospitals: Dana-Farber Cancer Institute, Brigham and Women’s Hospital, and Boston Children’s Hospital. N95 masks of the same model available in different sizes (for example, the 3M 1860 and the 3M 1860S, representing regular and small sizes) were considered to be interchangeable for testing purposes. The total sample size was necessarily limited by existing mask shortages and the importance of prioritizing the needs of healthcare workers; given the uniformity of the results reported below the sample size was judged to be adequate.

Additionally, an assortment of other PPE and hospital equipment was selected for sterilization. This included the following PAPR components: Sentinel^®^ XL CBRN hood with hose, Sentinel^®^ head cover hoods, Sentinel^®^ PAPR breathing tubes for use with Sentinel^®^ XL HP PAPR (ILC Dover, Frederica, DE), and Bullard RT Series PAPR hood (Bullard, Lexington, KY). Other equipment included two models of face shields, one locally fabricated^22^ and the other a Fisherbrand™ Disposable Face Shield (Fisher Scientific, Waltham MA), a DuPont Tyvek^®^ 400 coverall (Wilmington, DE) (**Supplementary Figure 1**), an iPad (Apple, Cupertino CA), and an iPad case. The iPad was included for testing since BWH is making them available to COVID-19 patients as a means of communicating with family members.

A first set of 30 N95 masks representing five different models was processed using the SteraMist system for zero to five cycles, and then analyzed for single-pass filtration efficiency using ambient particulate matter at MIT. A second set of 34 N95 masks was processed using the SteraMist system and sent to ICS Laboratories (Brunswick, OH) for testing to an abbreviated (instantaneous only) or full loading NIOSH N-95 filtration efficiency protocol. Nine masks underwent a quantitative fit test at DFCI following sterilization.

### Sterilization in a SteraMist environment chamber

Sterilization of N95 masks and other PPE was accomplished using a SteraMist-equipped room (dimensions 5.64 m × 4.57 m × 3.05 m) at the DFCI animal research facility. Three SteraMist environmental units (room version TPO-302-PLC-V1.4) are mounted on the ceiling of the room and can be controlled via a single panel, accessible from the outside. iHP mist was delivered through three nozzles at a total of 90 mL/min for 15 min, yielding a delivered concentration of 17.7 mL/m^3^.

N95 masks were placed with their interior surfaces facing up on standard stainless-steel shelves (open grid, InterMetro style). Most of the other PPE was also laid out on these shelves with the exception of two PAPR hoods, one PAPR hose, and the Tyvek coverall, which were hung in various configurations (**Figure 1**). PPE was spaced 6 cm to 20 cm apart on each shelf; this was designed to test sterilization performance at multiple points in the chamber. Tighter but non-overlapping spacing would likely be necessary for processing equipment in higher volumes. Two PAPR hoods, one PAPR hose, and one face shield were pre-treated with a SteraMist handheld spraying device in advance of processing in the SteraMist-equipped chamber. Pre-treatment was intended to ensure delivery of sterilant to items with semi-enclosed surfaces (such as the inside of a PAPR hose). Treating these items with the handheld device consisted of spraying the equipment from a distance of approximately 0.5–1m for a few seconds per surface. Per manufacturer protocol, the 100-minute sterilization cycle in the environmental chamber included: an initial 15-minute fill phase during which the mist was released; a 20-minute dwell phase to allow the mist to penetrate the room; and a 65-minute scrub phase during which the exhaust was re-opened to aerate the space at a rate of 43 air changes per hour. The room is tested quarterly to ensure homogeneous sterilization throughout the space.

**Figure 1:**
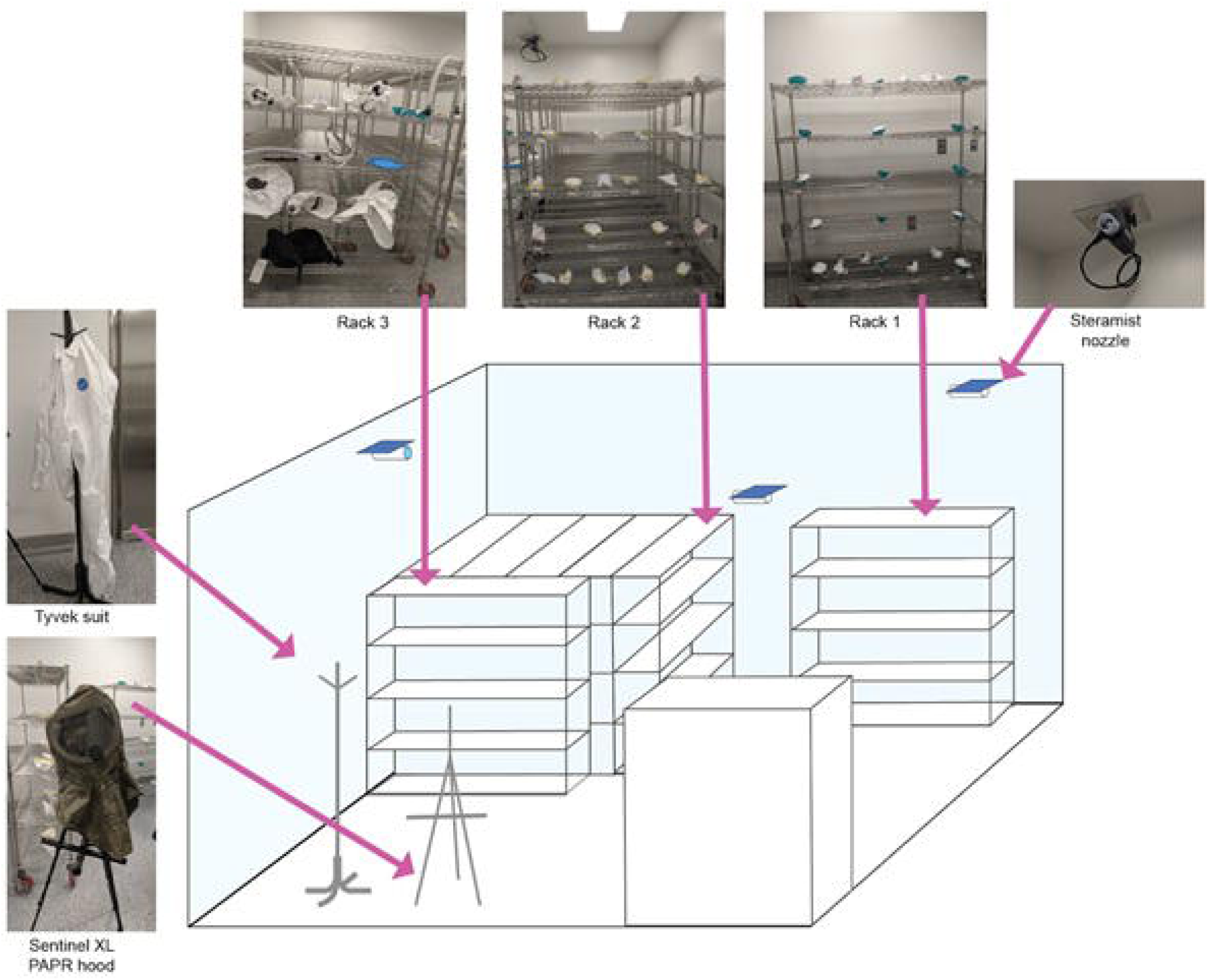
Diagram illustrating the testing set-up for SteraMist sterilization, with accompanying representative images. The chamber has an overall volume of ~80 m3.

### Evaluating sterility using biological indicators

The efficacy of sterilization was evaluated using Apex Biological Indicators (BIs: Mesa Labs; Boseman, MT); bacterial spores in these indicators are more resistant to killing than most viruses and therefore provide a conservative and simple estimate of sterilization efficacy. In particular, the *Geobacillus stearothermophilus* spores used in this study are known to be difficult to kill using hydrogen peroxide^6^. Each Apex biological indicator ribbon carries a minimum of 1.0 × 10^6^ *G. stearothermophilus* spores. The BIs were positioned in the environmental chamber prior to the first sterilization cycle. For N95 masks, BIs were placed under or adjacent to the masks. For the PAPR components and other equipment, BIs were placed on surfaces that were judged to be least accessible to the sterilant (for example, inside the PAPR tubing) (**Supplementary Table 1**). The BIs were extracted using sterile forceps, placed in Releasat growth medium (Mesa Labs), incubated at 55–60°C, and monitored for bacterial growth over a period of 10 days using a colorimetric assay^23^. Previous work with *G. stearothermophilus* spores suggests that a conservative benchmark for complete sterilization represents a 6-log_10_ kill; that is, a ratio in the number of surviving to initial viable spores in a BI of 10^−6^ (Reference ^24^). This corresponds to no observable bacterial growth and thus no color change after 5 days in Releasat medium.

### Evaluating filtration efficiency

Single-pass filtration efficiency testing was performed at MIT on five control N95 masks and 30 masks sterilized in the SteraMist-equipped chamber. An 8cm × 8cm sample of each N95 mask was inserted into a specialized air duct (**Supplementary Figure 2**), and ambient particulate matter was driven through the duct, and thus through the mask fabric, using a pressure differential of ~175 pascals at 0.4 m/s face velocity. The concentration of 0.3, 0.5, and 1 μm diameter particles prior to and after passage through the mask fabric was determined using an Aerotrak 9306 optical particle counter (TSI Inc.; Shoreview, MN) (**Table 2**). Filtration efficiency testing was performed a second time on a subset of decontaminated masks stored for 10 days after treatment to test for time-dependent degradation in N95 mask performance following sterilization. Although readily available and potentially effective, testing performed at MIT is not equivalent to NIOSH-approved testing for N95 masks and thus, these results should be interpreted as a relative, not absolute, measurements of filtration efficiency.

**Table 2:** Results obtained at a university laboratory on single-pass filtration efficiency for ambient particle matter. Each row represents a single N95 mask. Filtration efficiency values are an average of four upstream and downstream measurements.

| Model | Cycles | Filtration Efficiency (SD) |  |  |
| --- | --- | --- | --- | --- |
|  |  | 0.3µm | 0.5µm | 1.0µm |
| 3M 1860 | 0 | 97.66% (0.18) | 99.05% (0.13) | 99.68% (0.03) |
|  | 0 | 97.53% (0.18) | 99.11% (0.23) | 100.00% (0.00) |
|  | 1 | 99.20% (0.08) | 99.70% (0.08) | 99.90% (0.20) |
|  | 2 | 98.98% (0.10) | 99.80% (0.06) | 100.00% (0.00) |
|  | 2 | 99.42% (0.05) | 99.89% (0.09) | 99.91% (0.18) |
|  | 3 | 99.36% (0.12) | 99.92% (0.06) | 100.00% (0.00) |
|  | 4 | 98.55% (0.07) | 99.59% (0.13) | 99.88% (0.24) |
|  | 5 | 98.76% (0.03) | 99.52% (0.16) | 100.00% (0.00) |
|  | 10 | 98.45% (0.15) | 99.39% (0.09) | 100.00% (0.00) |
| KC/Halyard 46767 (duckbill) | 0 | 99.91% (0.02) | 99.95% (0.05) | 100.00% (0.00) |
|  | 1 | 99.83% (0.07) | 99.86% (0.17) | 100.00% (0.00) |
|  | 2 | 99.91% (0.02) | 99.98% (0.02) | 100.00% (0.00) |
|  | 3 | 99.90% (0.04) | 99.98% (0.04) | 100.00% (0.00) |
|  | 4 | 99.69% (0.06) | 99.80% (0.12) | 99.89% (0.24) |
|  | 5 | 99.89% (0.03) | 99.95% (0.07) | 100.00% (0.00) |
|  | 10 | 99.86% (0.07) | 99.97% (0.06) | 100.00% (0.00) |
| Gerson 2130 | 1 | 96.06% (0.20) | 98.90% (0.10) | 99.68% (0.43) |
|  | 2 | 96.46% (0.19) | 99.08% (0.11) | 99.84% (0.19) |
|  | 3 | 95.17% (0.37) | 98.80% (0.26) | 99.65% (0.30) |
| 3M 8210 | 0 | 98.09% (0.22) | 99.42% (0.24) | 99.82% (0.21) |
|  | 1 | 99.86% (0.04) | 99.99% (0.02) | 100.00% (0.00) |
|  | 2 | 99.52% (0.03) | 99.93% (0.04) | 100.00% (0.00) |
|  | 3 | 99.28% (0.06) | 99.88% (0.04) | 100.00% (0.00) |
|  | 4 | 98.90% (0.11) | 99.40% (0.10) | 100.00% (0.00) |
|  | 10 | 99.16% (0.15) | 99.77% (0.13) | 100.00% (0.00) |
| 3M 9210/37021 | 0 | 99.75% (0.11) | 99.92% (0.11) | 100.00% (0.00) |
|  | 1 | 99.77% (0.16) | 99.83% (0.19) | 99.71% (0.37) |
|  | 2 | 99.70% (0.07) | 99.92% (0.07) | 100.00% (0.00) |
|  | 3 | 99.39% (0.18) | 99.86% (0.04) | 100.00% (0.00) |
|  | 4 | 98.68% (0.98) | 99.01% (0.92) | 99.05% (1.19) |

A second sample of sterilized N95 masks was tested at ICS labs to NIOSH standards with 28 masks undergoing an instantaneous filter efficiency test and 6 masks undergoing a full loading filter efficiency test. Per NIOSH testing Procedure No. TEB-APR-STP-0059 (rev. 3.2)^25^, all masks were challenged with a sodium chloride aerosol neutralized to a Boltzmann equilibrium state at 25 ± 5 °C and a relative humidity of 30 ± 10%. Particle size and distribution was verified to correspond to a median diameter of 0.075 ± 0.020 µm, with a geometric standard deviation ≤ 1.86. N95 masks were conditioned at 85 ± 5% relative humidity and 38 ± 2 °C for 25 hours prior to filter efficiency testing. For instantaneous filter efficiency testing, each mask was then assembled into a fixture and subjected to instantaneous aerosol loading. The loading was performed by depositing sodium chloride aerosol at an airflow rate of 85 liters per minute (LPM) for one minute. For full loading filter efficiency, each mask was assembled into a fixture and subjected to full aerosol loading. The loading was performed by depositing 200 mg of sodium chloride aerosol at an airflow rate of 85 LPM for 75 minutes. Flow rate was monitored every 5–10 minutes on average and adjusted to maintain a flow rate of 85 ± 2 LPM.

### Quantitative fit testing

Nine masks from three models underwent a quantitative fit test following 2, 5, and 10 sterilization cycles to confirm that sterilization did not interfere with the ability of masks to form an effective seal with the human face. Testing was performed using a PortaCount Pro+ 8038 fit tester (TSI Inc.; Shoreview, MN) set to the 100-200 fit factor range, per manufacturer recommendation.

## Results

### Evaluating sterilization using biological indicators

All BIs placed under or adjacent to N95 masks that had been exposed to a single sterilization cycle in the SteraMist-equipped chamber exhibited at least a 9-log_10_ kill (representing no color change following seven days of incubation in Releasat medium). BIs placed within PAPR hoods also achieved 9-log_10_ kill as did a BI placed in a PAPR hose that was pre-treated using a SteraMist handheld spraying device (**Supplementary Table 1**). BIs placed on the iPad, iPad case, and PanFab face shield designs^22^ all passed the sterilization threshold, and the iPad was observed to be fully functional after one cycle of iHP treatment. In contrast, two BIs placed inside either end of a PAPR hose that was not subjected to pre-treatment were not sterilized, as determined by rapid bacterial growth following transfer to Releasat medium. This was also true of a BI embedded in the thick foam at the top of a Fisherbrand Disposable Face Shield. We tested the effect of pretreating the same face shield with a hand-held SteraMist device (after inserting a new BI) and observed a 4-log_10_ kill, which also fails the 6-log_10_ threshold conventionally used to score successful sterilization.

From these data we conclude that a single iHP cycle is efficacious at sterilizing N95 masks and other equipment placed throughout a SteraMist-equipped decontamination chamber and that the process is not obviously damaging to delicate equipment such as an iPad (n = 1). Penetration into semi-enclosed spaces such as PAPR hoses appeared to be less efficient, but such equipment could be sterilized by pre-treatment with a handheld iHP-delivery device followed by a cycle of iHP treatment in a chamber. Even with pre-treatment sterility was not achieved with thick face shield foam, suggesting that such normally disposable PPE should not be reused. In contrast, a custom-fabricated face shield^22^ introduced under an FDA EUA and consisting of 3D printed parts appeared to be sterilized effectively.

### Evaluating filtration efficiency

Performance data was collected at MIT on five models of N95 masks from three manufacturers (a total of 30 units) using an ambient particulate matter filtration efficiency test. Relative to control N95 masks, we observed no reduction in filtration efficiency for 0.3, 0.5, and 1 µm particles by N95 masks subjected to up to five sterilization cycles (**Table 2**). Data on pressure, temperature, air face velocity, and relative humidity during testing are found in **Supplementary Table 2**.

In addition, five models of N95 masks from three manufacturers (34 units total) were evaluated using testing protocols derived from NIOSH published standard testing procedures (STPs) maintained by ICS Laboratories. These data showed that 28 iHP-sterilized N95 masks retained instantaneous filtration efficiencies of ≥95%, including masks subjected to five sterilization cycles, the maximum number of cycles tested (**Figure 2, Table 3, Table 4, Supplementary Table 3, and Supplementary Material 1**). Gerson 2130 N95 masks were the least effective at filtering 75nm NaCl particles, but even these units passed the instantaneous test threshold out to five sterilization cycles. In no case did we detect an appreciable increase in resistance to airflow, a change which would likely be perceived by users as increased inhalation resistance.

**Figure 2:**
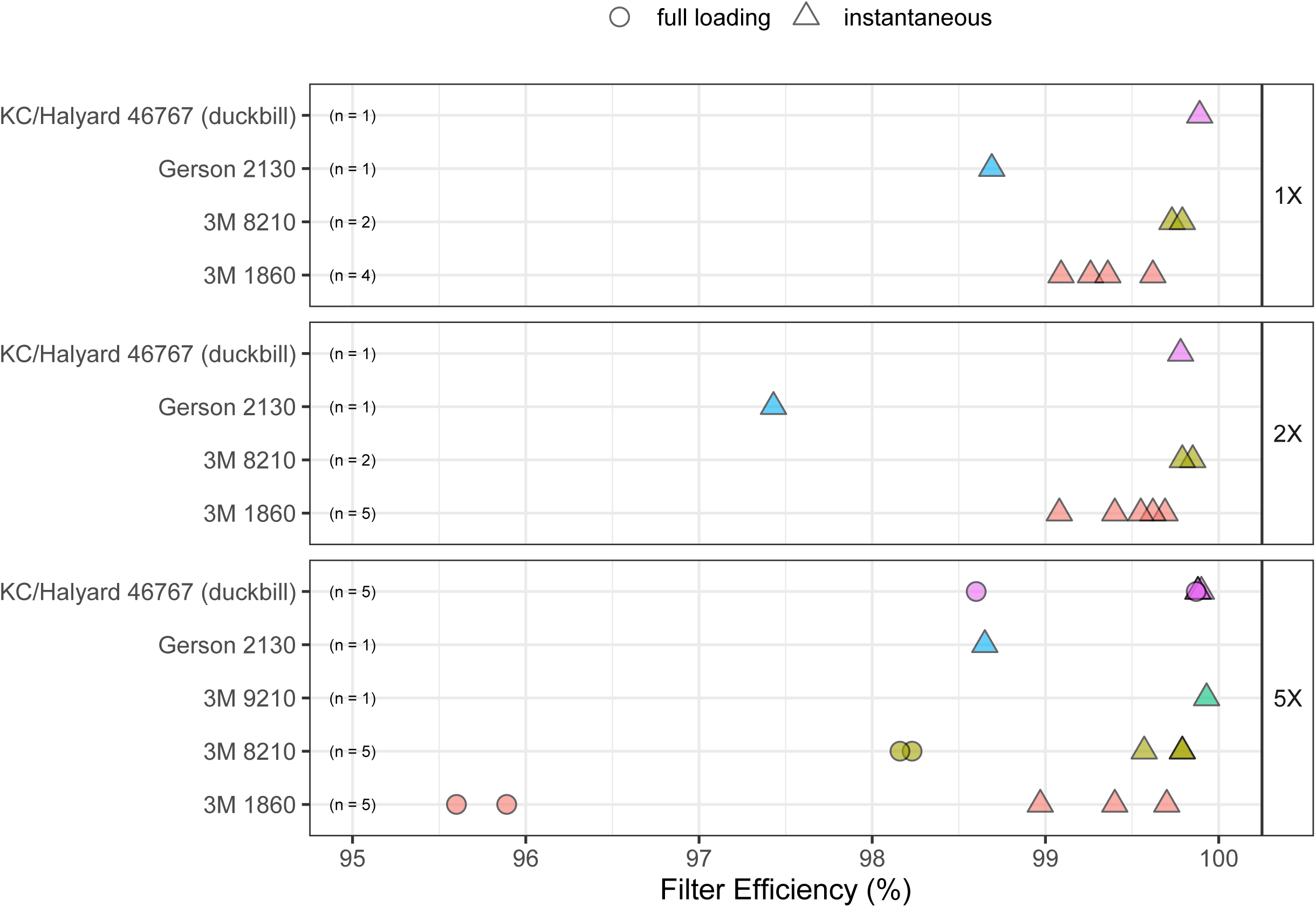
Instantaneous and fully-loaded ambient particulate matter filtration data for N95 masks over one, two, and five SteraMist sterilization cycles. Results were obtained from ICS Laboratories according to NIOSH standard Procedure No. TEB-APR-STP-0059. All masks passed ICS standards, including filtration efficiency of ≥ 95%.

**Table 3:** Results from ICS Laboratories on instantaneous filtration efficiency according to NIOSH standard Procedure No. TEB-APR-STP-0059. Each row represents data from 1–5 N95 masks and data are reported as the average for all tests that were performed.

| **Model** | **Cycles** | **Number of Masks** | **Flow Rate (LPM)** | **Resistance (mm of H<sub>2</sub>O)** | **Penetration (%)** | **Filter Efficiency (%)** | **Passed?** |
| --- | --- | --- | --- | --- | --- | --- | --- |
| **3M 1860** | 1 | 4 | 86 | 10.35 | 0.67 | 99.33 | Yes |
|  | 2 | 5 | 86 | 9.78 | 0.53 | 99.47 | Yes |
|  | 5 | 3 | 86 | 9.10 | 0.64 | 99.36 | Yes |
| **KC/Halyard 46767 (duckbill)** | 1 | 1 | 86 | 15.20 | 0.11 | 99.89 | Yes |
|  | 2 | 1 | 86 | 14.10 | 0.22 | 99.78 | Yes |
|  | 5 | 3 | 86 | 14.33 | 0.11 | 99.89 | Yes |
| **Gerson 2130** | 1 | 1 | 86 | 9.30 | 1.31 | 98.69 | Yes |
|  | 2 | 1 | 86 | 7.90 | 2.57 | 97.43 | Yes |
|  | 5 | 1 | 85 | 9.80 | 1.35 | 98.65 | Yes |
| **3M 8210** | 1 | 2 | 86 | 8.85 | 0.24 | 99.76 | Yes |
|  | 2 | 2 | 86 | 8.95 | 0.18 | 99.82 | Yes |
|  | 5 | 3 | 85 | 9.40 | 0.28 | 99.72 | Yes |
| **3M 9210** | 5 | 1 | 85 | 10.40 | 0.07 | 99.93 | Yes |

**Table 4:** Results from ICS Laboratories on full loading filtration efficiency according to NIOSH standard Procedure No. TEB-APR-STP-0059. Each row represents data from an N95 mask.

| Model | Cycles | Flow Rate (LPM) | Resistance (mm of H<sub>2</sub>O) | Initial Penetration (%) | Maximum Penetration (%) | Filter Efficiency (%) | Passed? |
| --- | --- | --- | --- | --- | --- | --- | --- |
| 3M 1860 | 5 | 85 | 9.6 | 1.22 | 4.40 | 95.60 | Yes |
|  | 5 | 85 | 9.8 | 0.94 | 4.11 | 95.89 | Yes |
| KC/Halyard 46767 (duckbill) | 5 | 85 | 15.5 | 1.40 | 1.40 | 98.60 | Yes |
|  | 5 | 85 | 14.8 | 0.13 | 0.13 | 99.87 | Yes |
| 3M 8210 | 5 | 85 | 9.8 | 0.29 | 1.77 | 98.23 | Yes |
|  | 5 | 85 | 9.9 | 0.49 | 1.84 | 98.16 | Yes |

Fully loaded filtration efficiency was also evaluated by ICS Laboratories to NIOSH standards. The purpose of this test is to mimic the effect of an accumulation of charged particles in a mask, a phenomenon related to time of use. Mask loading is known to reduce filtration efficiency, potentially by weakening electrostatic charge in the filtering layer^26^. Again, we observed that the 9 sterilized masks passed the NIOSH threshold for N95 pre-certification.

To test for time-dependent degradation of performance, 26 N95 masks were tested at MIT 10 days after initial filtration testing **(Supplementary Table 4)** and 10-15 days after sterilization with iHP. We observed no difference in filtration performance between measurements taken immediately after sterilization, and those taken 10 days after, as determined by a repeated measures ANOVA (p = 0.45). From these data we conclude that the filtration efficiency of multiple models of domestically manufactured N95 masks is not substantially affected by one to five cycles of iHP sterilization in terms of filtration efficiency or inhalation resistance, and that all masks tested meet existing NIOSH pre-certification standards.

### PortaCount quantitative fit data

All nine masks that were tested for fit (KC/Halyard 46767, 3M 1860, 3M 8210) using the PortaCount equipment passed a reading of >200 fit factor following 2, 5, and 10 sterilization cycles. This corresponds to a filtration efficiency of 99% or higher (data not shown) according to manufacturer guidelines. Thus, iHP sterilization does not appear to impair the ability of N95 masks to form an effective seal against a user’s face.

## Discussion

Hydrogen peroxide has a long history of successful use in the field of medical device sterilization, and our results support the use of iHP as a PPE sterilant when delivered using a SteraMist-equipped environment chamber, in some cases complemented by pre-treatment with a handheld iHP delivery device. Thus, iHP sterilization can likely be used to extend the usability of PPE such as N95 masks that are usually disposed of after a single use. The DFCI SteraMist environment chamber used in this study has a volume of ~80 m^3^ and could comfortably fit ~ 2400 N95 masks per cycle without the masks touching each other, or a lesser number of PAPR hoods and other PPE. At this rate, assuming idealized staffing and logistics, roughly 4,800-7,200 masks could be sterilized for use per day given a typical 100 minute sterilization cycle. These numbers could be increased with the addition of an overnight workforce.

In keeping with standard practice, sterility was judged in this study using biological indicators containing bacterial spores and was not based on killing of pathogens such as SARS-CoV-2 encountered in a clinical setting. However, the *G. stearothermophilus* spores in the BIs we used are known to be resistant to killing by hydrogen peroxide, and spores are substantially more resistant to sterilization than enveloped viruses such as SARS-CoV-2^27^. Recent work has also demonstrated that VHP can kill SARS-CoV-2.^28^ Thus, we do not believe our use of BIs rather than direct measurement of viral viability represents a significant limitation in the interpretation of the data.

Two independent lines of evidence, one generated at a university laboratory and one at a commercial laboratory accredited to perform N95 mask certification to NIOSH/ISO/IEC standards, show that N95 masks decontaminated with iHP using SteraMist technology retain their performance with respect to filtration and inhalation resistance for at least five cycles, the maximum number tested. No deterioration was detected in masks tested 10 days post treatment. Quantitative fit testing of sterilized N95 masks confirms that they still form an airtight seal as required (in this case, tests were performed out to 10 cycles). Thus, sterilized N95 masks remain fully functional.

### Limitations of this study

Given the urgency of N95 mask shortages, we are reporting results obtained to date. However, some additional tests are currently underway to increase confidence in our specific findings and the practice of sterilizing and reusing N95 masks more generally. For example, while we report complete data on testing for up to 5 cycles on N95 masks, further evaluation of masks that have undergone 10 cycles of sterilization are underway at ICS Labs, using both instantaneous and fully loaded filtration testing. The tests described in this study were conducted using unused N95 masks. We do not yet have data on N95 masks used in an actual health care facility responding to a pandemic. Among questions to be addressed by real-world testing are inhalation resistance for an N95 mask that has been loaded with internal and external contaminants, the comfort level of health care workers in using an N95 mask that is sterilized but previously used by another individual, and the rate of wastage arising from breakage of elastic bands, contamination with makeup or topical face products, and unacceptable degradation in fit. Remarkably, these types of real-world use data are not available for any iHP/VHP-based sterilization method, even for technologies that have been heavily promoted commercially. Nonetheless, the data reported in this study were judged by our clinical teams to be sufficient to implement N95 mask sterilization and reuse at DFCI.

Future work should address the question of whether decontaminated N95 masks must be returned to the original users (as specified in the Sterrad and Sterris EUAs for N95 mask decontamination) or can be returned to a common pool (as specified in the Batelle EUA); the latter is substantially easier to implement from a logistical perspective. Finally, while there is good support for the use of spore-based BIs in measuring the efficacy of sterilization, direct tests on SARS-CoV-2 itself may be warranted, particularly in the case of items such as hoses and other PPE that have a complex shape (this would require BL3-level studies).

## Conclusions

Our data support the use of the SteraMist iHP technology as a sterilization method for reuse of N95 masks, including many of the most commonly used models, as well as other types of PPE – in some cases following pre-treatment with an iHP handheld delivery device. In interpreting these data, it is important to note that not all iHP/VHP methods are the same. While Bioquell is approved under an FDA EUA for 20 cycles, N95 masks sterilized using an alternative HPGP method commercialized by Sterrad fail at five cycles (the Sterrad EUA was approved for 2 sterilization cycles and requires that a mask be returned to a single user).^8,29^ Moreover, our data show that semi-enclosed items of PPE, such as PAPR hoses, cannot be sterilized without pre-treatment, and that face shields with thick foam may not be sterile even after iHP pre-treatment followed by iHP treatment in an environmental chamber. Thus, it is imperative that institutions seeking to deploy iHP/VHP technology review available primary data prior to local deployment. We also suggest that BIs routinely be deployed when attempting to sterilize hoses and other semi-enclosed PE components.

The issues described above about reuse of N95 masks have been recognized for over two decades based on multiple instances of human transmission of novel respiratory diseases. As the global response to COVID-19 evolves, we hope that the study of sterilization technologies such as iHP/VHP will continue and involve peer-review of independently acquired data so that we are in a better position for the coming waves of this crisis and for those in the future.

## Data Availability

The datasets generated during and/or analysed during the current study are available as supplementary materials.

## Acknowledgments

Above all we thank the members of the Greater Boston Pandemic Fabrication Team (PanFab) for technical, administrative, and logistical support necessary for the execution of this project. Membership found at https://www.panfab.org/the-team-and-the-project/consortium-members. We also thank Prof. Mike Short (MIT Nuclear science and Engineering); Karen Byers (DFCI Biosafety); Chad Pires (Boston Children’s Hospital Environmental Safety); and Dale Pfriem along with the employees of ICS Laboratories for performing N95 mask testing under challenging conditions.

## Author Contributions

Study conception and planning: A.K.C, D.P., H.Y, M.C., F.T.B, R.G., S.H.Y., P.K.S, N.R.L.

Study execution: A.K.C., H.Y., E.T, D.K., D.T., J.L., H.M.

Writing: A.K.C, D.P., H.Y, M.C., M.S.S., F.T.B, J.L, R.G., S.H.Y., P.K.S, N.R.L.

Greater Boston Pandemic Fabrication Team Coordination: D.P., H.Y., P.K.S.

Guarantors: S.H.Y., P.K.S, N.R.L.

## Public and Patient Involvement statement

Patient and members of the public were not involved in this study.

## Dissemination declaration

Dissemination of results to study participants and or patient organizations is not applicable.

## Ethics approval

Ethics approval was not necessary to conduct this study.

## Transparency statement

S.H.Y., P.K.S, N.R.L. affirm that the manuscript is an honest, accurate, and transparent account of the study being reported; that no important aspects of the study have been omitted; and that any discrepancies from the study as originally planned have been explained.

## Data sharing

All data relevant to the study are included in the article or uploaded as supplementary information.

## Supplementary Material Legends

**Supplementary Figure 1:** Illustrative photograph of handheld device processing before sterilization by environmental SteraMist system, on a DuPont Tyvek® 400 coverall.

**Supplementary Figure 2:** Image of ambient particulate matter air ducts used in MIT testing of N95 masks. Note that this is a destructive test in which the N95 mask is cut prior to introduction into the testing chamber.

**Supplementary Table 1:** Results of biological indicator (BI) monitoring after one-cycle of sterilization in a SteraMist-equipped environment chamber.

**Supplementary Table 2:** Results of ambient particulate matter filtration efficiency evaluation performed at MIT, recorded within 24 hours of treatment. Values are an average of four upstream and downstream measurements, with standard deviation (SD) shown.

**Supplementary Table 3:** Results of instantaneous filtration efficiency evaluation performed at ICS Laboratories (Brunswick, OH).

**Supplementary Table 4:** Results of ambient particulate matter filtration efficiency evaluation re-performed at MIT 10 days after sterilization treatment. Values are an average of four upstream and downstream measurements.

**Supplementary Material 1:** Official report with results from all testing performed at ICS Laboratories.

#### SUMMARY BOX

##### Section 1: What is already known on this topic

- The COVID-19 pandemic has led to widespread shortages in personal protective equipment, such as N95 masks, and there have been corresponding attempts for mask sterilization and subsequent reuse.
- There is an absence of publicly available data on the post-sterilization performance of different makes and models of N95 masks following different sterilization or decontamination procedures.

##### Section 2: What this study adds

- This study evaluates a recently developed technology, ionized hydrogen peroxide (iHP), as a method for sterilizing N95 masks and other PPE. Specifically, it assesses the SteraMist Binary Ionization Technology® (BIT) from TOMI installed at the Dana-Farber Cancer Institute (DFCI; Boston, MA) animal research facility.
- Masks were tested for three critical features after multiple sterilization cycles: (1) sterility, as measured by the inactivation of bacterial spores contained in biological indicators; (2) filtration efficiency, measured both by aerosolized 75 nm NaCl particles and by 0.3-1 µm ambient particulate matter; and (3) fit, using a PortaCount quantitative fit test apparatus. Sterilization of other PPE items such as face shields as well as hoods and hoses for Powered-Air Purifying Respirators (PAPRs) was also explored.
- Our data support the use of the SteraMist iHP technology as a sterilization method for reuse of N95 masks, including many of the most commonly used models, as well as other types of PPE.

